# The Potential Clinical and Economic Impact of the Next-Generation COVID-19 mRNA-1283 Vaccine in Canada

**DOI:** 10.1101/2025.05.29.25328577

**Authors:** Kelly Fust, Michele Kohli, Shannon Cartier, Keya Joshi, Nicolas Van de Velde, Ekkehard Beck, Michelle Blake

## Abstract

**Background:** With continued high disease burden observed in vulnerable groups and fiscal responsibility shifting to Canada’s jurisdictions, assessing the economic value of COVID-19 vaccines is critical for optimizing COVID-19 prevention. This study estimated the public health impact and economically justifiable price (EJP) of Moderna’s next-generation COVID-19 vaccine (mRNA-1283) versus no vaccination in Canada, and relative to currently authorized COVID-19 vaccines (mRNA-1273; BNT-162b2).

**Methods:** The target population included individuals aged ≥65 years and 12-64 years at high-risk of severe COVID-19 outcomes, consistent with 2025/2026 national guidelines. Analyses were conducted using a static decision-analytic model (1-year horizon) from a publicly funded healthcare payer perspective. Vaccine efficacy against infection and hospitalization for mRNA-1283 versus no 2024/2025 vaccination was based on mRNA-1283’s pivotal trial and mRNA-1273 real-world data. Clinical outcomes included symptomatic infections, hospitalizations, deaths, and number needed to vaccinate (NNV); economic outcomes included total costs, quality-adjusted life-years (QALY), and EJP at a $50,000/QALY willingness-to-pay threshold. Sensitivity analyses were performed.

**Results:** Compared to no vaccine, annual vaccination with mRNA-1283 prevented 288,912 symptomatic infections (NNV=15), 11,710 hospitalizations (NNV=364), and 2,194 deaths (NNV=1,944). The EJP for mRNA-1283 was $325 ($230-$771 in scenario analyses). Semi-annual dosing for those ≥65 years or ≥80 years averted additional hospitalizations and deaths compared to annual vaccination. mRNA-1283 prevented an additional 2,873-3,689 hospitalizations and 537-690 deaths compared to currently authorized vaccines. EJPs for mRNA-1283 were $78 and $103 when compared to mRNA-1273 and BNT162b2, respectively.

**Interpretation:** mRNA-1283 could reduce the COVID-19 clinical burden and provide economic value for the NACI-recommended population, exceeding current COVID-19 mRNA vaccines.

## Introduction

In the post-pandemic era, COVID-19 remains a persistent public health challenge in Canada, with multiple peaks occurring year-round.(1-5) COVID-19 continues to cause more hospitalizations and deaths than influenza, yet vaccine coverage remains lower, and the burden disproportionately affects older adults aged ≥65 years and those with underlying medical conditions.(1-5) Despite strong recommendations from the National Advisory Committee on Immunization (NACI) emphasizing the effectiveness and safety of COVID-19 vaccines for these vulnerable groups, protection for these individuals remains suboptimal.(4, 6, 7) Additionally, with the integration of COVID-19 vaccination into jurisdictional immunization programs, provinces and territories now bear the fiscal responsibility for vaccine procurement and delivery.(3, 4) Thus, policy-makers face the challenge of optimizing COVID-19 vaccination strategies to most effectively and efficiently protect those vulnerable to severe COVID-19 disease. Alongside considerations of clinical benefit-risk and other factors, the assessment of health economic value of COVID-19 vaccination—both overall and by vaccine product—can help guide resource allocation and support priority-setting across jurisdictions.(8)

mRNA-1283 is an investigational, next-generation COVID-19 vaccine engineered to generate a more targeted immune response. In the pivotal Phase 3 NextCOVE (P301) randomized clinical trial, mRNA-1283 exhibited a comparable safety profile and stronger immune response compared with the currently authorized mRNA-1273 vaccine.(9-11) To date, neither the comparative economic value of mRNA-1283 nor a publicly available list price for any COVID-19 vaccine has been established in Canada. Using data on relative vaccine efficacy (rVE) and safety data from the pivotal mRNA-1283 trial, alongside Canadian-specific burden of disease, costs and resource use inputs, this study therefore aimed to estimate both the overall health impact and economically justifiable price (EJP) for mRNA-1283 compared to no vaccination (i.e. no annual or semi-annual COVID-19 vaccination in the 1-year time horizon) based on Canada’s commonly used willingness-to-pay (WTP) threshold of $50,000 per quality-adjusted life-year (QALY) gained.(12-16) A secondary objective was to estimate the health impact and EJP relative to currently licensed mRNA vaccines (mRNA-1273, BNT-162b2). Accordingly, we aim to inform vaccine choice within Canada’s evolving immunization landscape—and support a more resilient, efficient response to COVID-19.

## Methods

### Analyses

Analyses were conducted with a static decision-analytic model across a 1-year time horizon starting in September. The base-case target population included individuals aged ≥65 years and 12-64 years at high-risk of severe COVID-19 outcomes in Canada, with a single annual dose assumed, consistent with the 2025/2026 National Advisory Committee on Immunization (NACI) guidelines(3,4) and the mRNA-1283 expected authorization for ages ≥12 years. Scenario analyses examined the population most likely to benefit clinically(3) from an annual dose of mRNA-1283 (adults ≥65 years and high-risk individuals 18-64 years); and the impact of a second dose (semi-annual dosing) administered to groups identified as key candidates within NACI’s post-pandemic recommendations(3,4): 1) ≥65 years and 2) ≥80 years. Annual dosing was assumed to begin in October of 2024. When semi-annual dosing was considered, second doses started in March of the following year and a minimum of 3-months between doses was assumed.

Clinical outcomes of the base-case, scenario and deterministic sensitivity analyses (DSAs) included COVID-19 symptomatic infections, hospitalizations, deaths, and cases of ‘Long COVID’/ post-COVID-19 condition (PCC). Economic outcomes included total costs, total quality-adjusted life-years (QALYs) lost, and the EJP (including 10% vaccine wastage(17,18)) at a $50,000/QALY gained WTP threshold(12,15,16,19,20) (see EJP calculations in the Technical Appendix). Lifetime QALY losses associated with premature death due to COVID-19 infection during the one-year time horizon, discounted to prevent value at a rate of 1.5%,(21) were also included. Base-case analyses were performed from the publicly funded healthcare payer perspective with a scenario analysis from the societal cost perspective.

### Model Structure

The model followed the target population on a monthly basis and moved the proportion that developed COVID-19 symptomatic infection through a decision-tree(22) (Figure 1), which calculated numbers of hospitalizations and deaths, and associated costs and QALYs lost. The monthly uptake of COVID-19 vaccine was tracked and the number of adverse events experienced and associated costs and QALYs lost were estimated. Vaccination effectiveness (VE) reduced the probability of infection and could further reduce the probability of hospitalization following infection. Initial VE was assigned in the first month that a vaccine was given and then declined linearly on a monthly basis; the model tracked the VE over time. The model calculated the difference between hospitalization and infection VE and reduced the probability of hospitalization following infection by the resulting incremental VE (See Technical Appendix).

**Figure 1.**
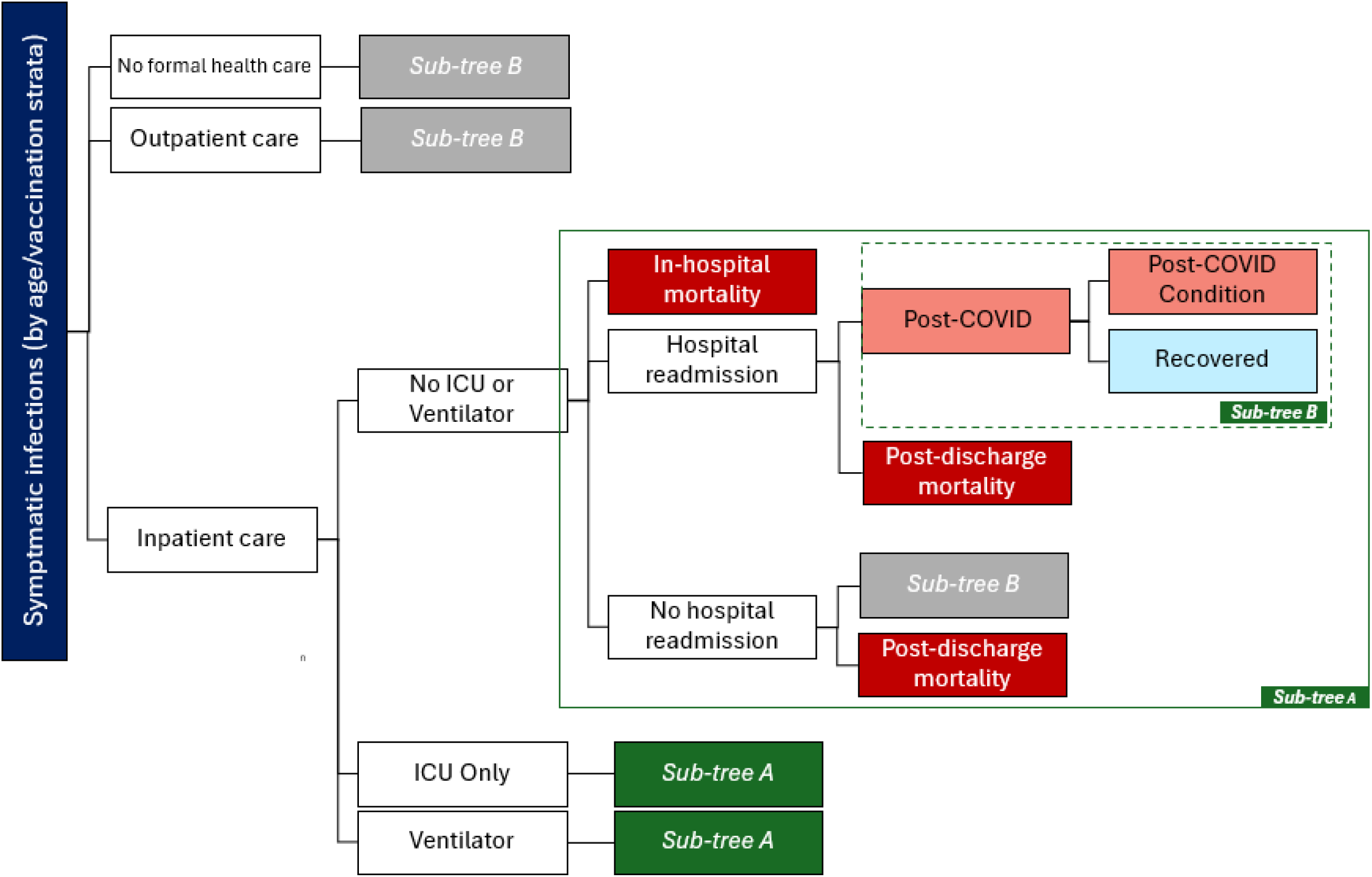
Model Structure. ICU: Intensive Care Unit -Not shown in figure: all symptomatic infections are at risk of infection-related myocarditis, and all hospitalizations are followed by post-hospital recovery. An average cost and QALY loss may be assigned for each of these events. -Ventilator refers to mechanical ventilation in an intensive care setting.

### Infection Incidence

The model required the monthly incidence of symptomatic SARS-CoV-2 infection in the Canadian population that did not receive an annual vaccine. As these data are no longer published by any Canadian jurisdiction, an estimation process was developed using age-specific hospitalization rates per 100,000. As the hospitalization data reflect a partially vaccinated population, information on vaccination coverage, VE, and the probability of hospitalization must also be used in this estimation process (Technical Appendix). Given the uncertainty of post-pandemic COVID-19 incidence, four different estimates of monthly hospitalizations (Technical Appendix) were used. The base-case (incidence 1) and an alternative scenario (incidence 2) were based on projections provided by Miranda et al. (2024).(17) Additional scenarios for British Columbia (BC)(23) and Quebec(24) were developed using hospitalization data from the 2023-2024 season.

### Vaccine Effectiveness

All derived initial 2024-2025 VE values are displayed in Table 1 and described further in the Technical Appendix. The VEs of mRNA-1273 targeting the KP.2 variant for those ≥18 years were based on a real-world evidence study that estimated the VE against medically-attended COVID-19 (used as a proxy for infections) and COVID-19 related hospitalizations.(25,26) For those aged 12-17 years, due to the lack of updated and disaggregated data in this age group, the initial 2024-2025 VEs for mRNA-1273 against infection and hospitalization were assumed to be the same and based on the VE of XBB.1.5 vaccines against emergency department and urgent care visits.(27) Relative VE (rVE) data for mRNA-1283 compared to mRNA-1273 from the NextCOVE phase 3 pivotal study (Study P301) were used to obtain the 2024-2025 VE for mRNA-1283. BNT-162b2 was calculated using the rVE of mRNA-1283 compared to BNT-162b2 from an indirect treatment comparison.(28,29) Table 2 shows the inputs used for DSAs comparing mRNA-1283 and no annual vaccination.

**Table 1.** Initial vaccine effectiveness values (%) for all vaccines considered in the analyses.

|  | mRNA-1273 | rVE of mRNA-1283 compared to mRNA-1273 | mRNA-1283 | rVE of mRNA-1283 compared to BNT-162 | BNT-162b2 |
| --- | --- | --- | --- | --- | --- |
| **Symptomatic Infection** |  |  |  |  |  |
| **12 -17 years (high-risk)** | 71.0 | 17.5 | 76.1 | -- | 71.0* |
| **18 - 64 years (high-risk)** | 48.2 | 17.5 | 57.3 | 20.5 | 46.2 |
| **≥ 65 years** | 54.5 | 13.5 | 60.7 | 22.8 | 49.0 |
| **Hospitalization** |  |  |  |  |  |
| **12 -17 years (high-risk)** | 71.0 | 38.1 | 82.0 | -- | 71.0* |
| **18 - 64 years (high-risk)** | 50.6 | 38.1 | 69.4 | 44.1 | 45.2<sup>‡</sup> |
| **≥ 65 years** | 57.2 | 38.1 | 73.5 | 44.1 | 52.6 |

rVE: relative VE
\* Assumed to be the same as mRNA-1273
‡ The model uses a different VE estimate (VE against infection: 46.2%), as the model requires VE against infection <= VE against hospitalization.

**Table 2.** Comparison to no vaccine: initial vaccine effectiveness values (%) for mRNA-1383, base-case and sensitivity analyses.

|  | **Base Case\*** | **Lower 95% CI** | **Upper 95% CI** | **Scenario: rVE against infections equal to rVE against hospitalization** |
| --- | --- | --- | --- | --- |
| Infection |  |  |  |  |
| 12 -17 years (high-risk) | 76.1 | 59.8 | 91.5 | 76.1 |
| 18 - 64 years (high-risk) | 57.3 | 49.1 | 65.0 | 57.3 |
| ≥ 65 years | 60.7 | 50.8 | 70.0 | 60.7 |
| Hospitalization |  |  |  |  |
| 12 -17 years (high-risk) | 82.0 | 64.2 | 95.3 | 76.1 |
| 18 - 64 years (high-risk) | 69.4 | 46.3 | 88.2 | 59.2 |
| ≥ 65 years | 73.5 | 53.3 | 89.9 | 63.0 |

CI: Confidence interval; rVE: relative vaccine effectiveness

The absolute monthly waning rate was assumed to be the same for all vaccines: 4.75% against infection(30) and 2.46% against hospitalization.(31) Ranges based on 95% CI of 3.05%-6.75%(30) for infection and 1.37%(30) to 3.87%(32) for hospitalization were used in DSAs.

### Other model inputs

Parameter values for the base-case, scenario, and deterministic sensitivity analyses (probabilities, costs, and QALYs) are detailed in the Technical Appendix. Optimal timing of vaccine administration for the base case population was examined in scenario analyses by shifting the start of vaccination from October to September and November.

## Results

Clinical outcomes for the base-case and target population scenario analyses are presented in Table 3. Compared to no vaccine, vaccinating all ≥65 years and high-risk 12-to-64-year-olds annually would prevent 288,912 symptomatic infections, 11,710 hospitalizations, and 2,194 deaths. The NNVs to prevent one infection, hospitalization, or death are 15, 364, and 1,944, respectively. If all Canadians aged ≥80 years were offered a second dose, mRNA-1283 would prevent an additional 1,483 symptomatic infections, 54 hospitalizations, and 11 deaths. Decreasing the age threshold for a second dose from ≥80 years to ≥65 years would prevent an additional 5,544 symptomatic infections, 100 hospitalizations, and 19 deaths compared to annual dosing.

**Table 3.** Clinical Outcomes, by Target Population.

| Target Population | Vaccination Strategy | Outcomes |  |  |  | Outcomes Averted<br>(Compared to no vaccine) |  |  |  |
| --- | --- | --- | --- | --- | --- | --- | --- | --- | --- |
|  |  | Infections | Hosp. | PCC | Deaths | Infections | Hosp. | PCC | Deaths |
| *Base-case (65+ all; 12-64 HR annual dosing)* | No vaccination | 3,244,588 | 53,913 | 45,627 | 9,744 | -- | -- | -- | -- |
|  | mRNA-1283 | 2,955,676 | 42,204 | 39,521 | 7,551 | 288,912 | 11,710 | 6,106 | 2,194 |
| 65+ all; 12-64 HR(65+ semi-annual dosing) | mRNA-1283 | 2,950,131 | 42,104 | 39,384 | 7,532 | 294,456 | 11,809 | 6,243 | 2,212 |
| 65+ all; 12-64 HR(80+ semi-annual dosing) | mRNA-1283 | 2,954,789 | 42,171 | 39,469 | 7,544 | 289,799 | 11,742 | 6,158 | 2,200 |
| 18-64 High-Risk, All Adults 65+ (Annual Dosing) |  |  |  |  |  |  |  |  |  |
| 65+ all; 18-64 HR(annual dosing) | No vaccination | 3,149,068 | 53,860 | 45,627 | 9,741 | -- | -- | -- | -- |
|  | mRNA-1283 | 2,861,246 | 42,151 | 39,521 | 7,547 | 287,821 | 11,709 | 6,106 | 2,193 |

HR: High-risk; Hosp.: Hospitalizations; PCC: post-COVID condition (long COVID)
65+ all; 12-64 HR: Includes the population aged 12-64 years of age at high-risk, and the general population ≥65 years of age.
65+ all; 18-64 HR: Includes the population aged 18-64 years of age at high-risk, and the general population ≥65 years of age.

Excluding high-risk 12-to-17-year-olds, thus narrowing annual vaccination to all ≥65 years and high-risk 18-to-64-year-olds, would prevent 287,821 symptomatic infections, 11,709 hospitalizations, and 2,193 deaths relative to no vaccine. The NNVs to prevent one infection, hospitalization, and death are 15, 363, and 1,938, respectively.

The EJP for the base-case analysis at a $50,000/QALY WTP threshold was $325 versus no vaccine. Expanding the vaccination strategy to include semi-annual dosing for adults aged ≥80 and ≥65 years would yield EJPs of $297 and $258, respectively, compared to no vaccine. Vaccinating a population of all ≥65 years and high-risk 18-64-year-olds would yield an EJP of $326 compared to no vaccine.

Results of DSAs on hospitalizations and deaths prevented by mRNA-1283 compared to no vaccination are presented in Figure 2. Results are most sensitive to variations in COVID-19 incidence. Using incidence estimates based on Quebec data yields 32,072 hospitalizations prevented by mRNA-1283 (174% increase from base-case) and 6,317 deaths prevented (188% increase); using BC data yields 17,270 hospitalizations (47% increase) and 3,413 deaths prevented (56% increase). Model results are also sensitive to mRNA-1283 initial VE for hospitalization, with hospitalizations prevented ranging from 14,605 to 9,451 when the VE is varied to the 95% CI upper and lower bounds, respectively. Deaths prevented are sensitive to in-hospital mortality estimates, ranging from 1,753 to 2,634 deaths prevented when estimates are varied ±25% from the base-case. Shifting estimates of vaccine uptake from October to start one month later (November) results in 2,250 fewer hospitalizations and 421 deaths prevented compared to the base-case, respectively; shifting earlier (September) would prevent an additional 1,706 hospitalizations and 318 deaths compared to the base-case. The impact of various incidence scenarios on hospitalization rates is presented in Figure 4; the impact of shifting vaccination timing to start in either September or November on hospitalizations prevented by mRNA-1283 is displayed in Figure 5.

**Figure 2.**
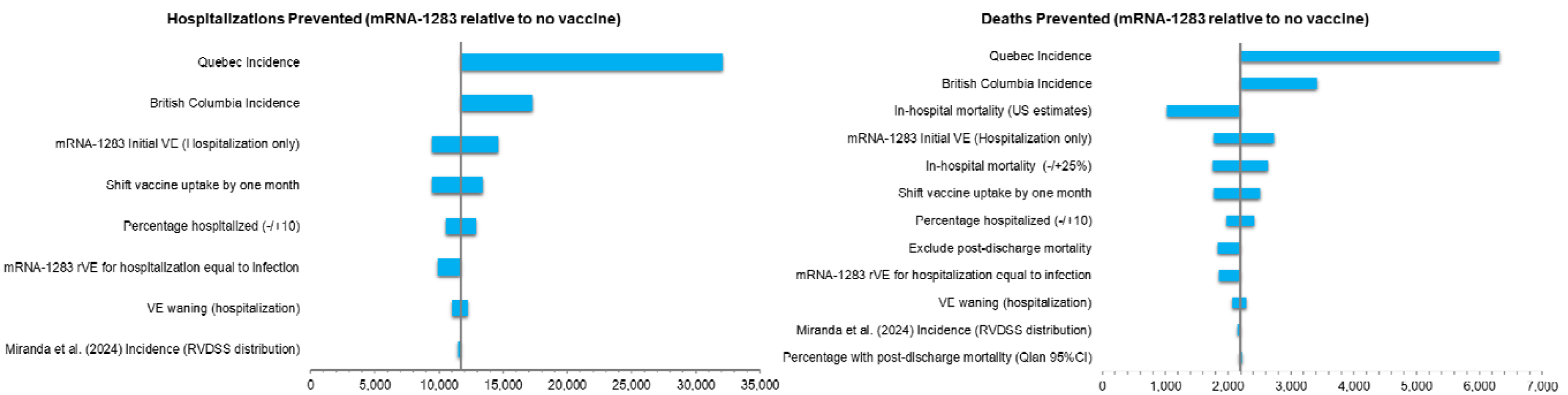
Clinical Sensitivity Analyses (65+ All, 12-64 High-Risk, with Annual Dosing) CI: Confidence Interval; RVDSS: Respiratory Virus Detection Surveillance System; rVE: relative vaccine effectiveness; US: United States; VE: Vaccine effectiveness

Figure 3 shows the range of EJPs for mRNA-1283 versus no vaccine, with a base-case EJP of $325 at a $50,000/QALY threshold, ranging from $230 (−29%) using U.S. in-hospital mortality data to $771 (+137%) using Quebec incidence estimates.

**Figure 3.**
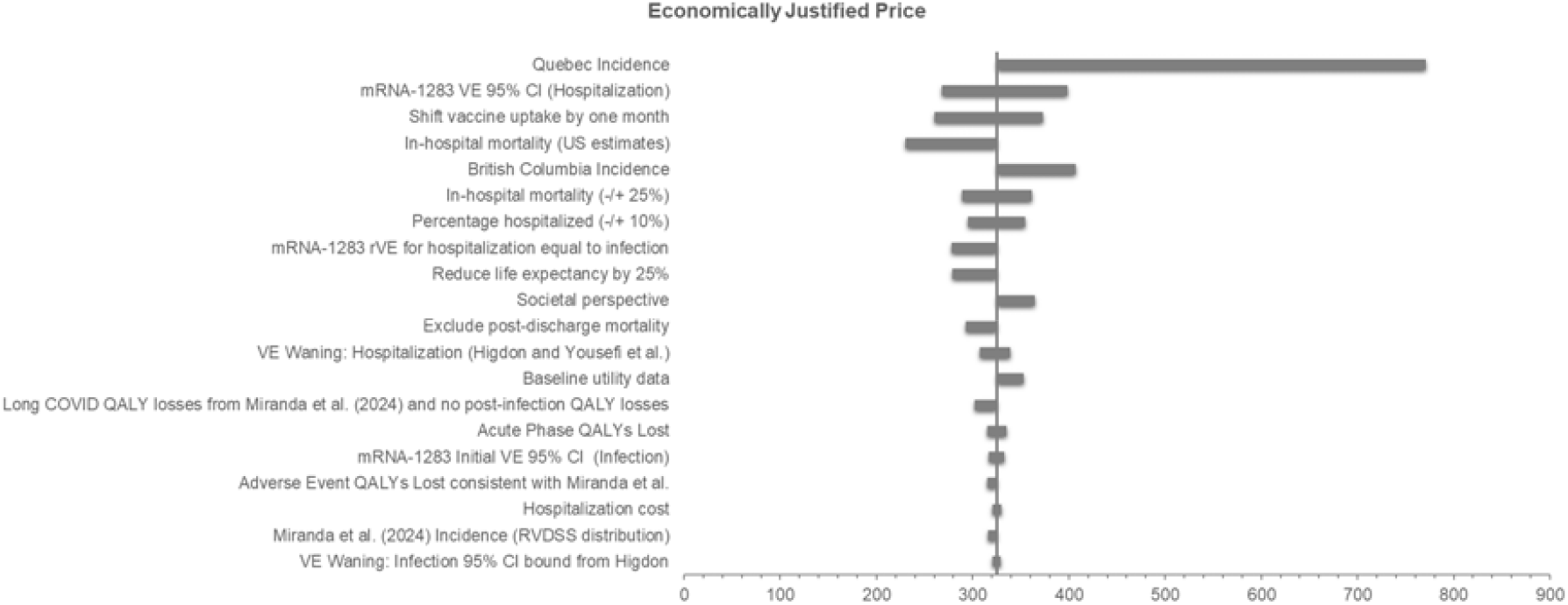
mRNA-1283 EJP at $50,000 per QALY WTP Threshold (65+ All, 12-64 High-Risk, with Annual Dosing) CI: Confidence Interval; RVDSS: Respiratory Virus Detection Surveillance System; rVE: relative vaccine effectiveness; US: United States; VE: vaccine effectiveness

**Figure 4.**
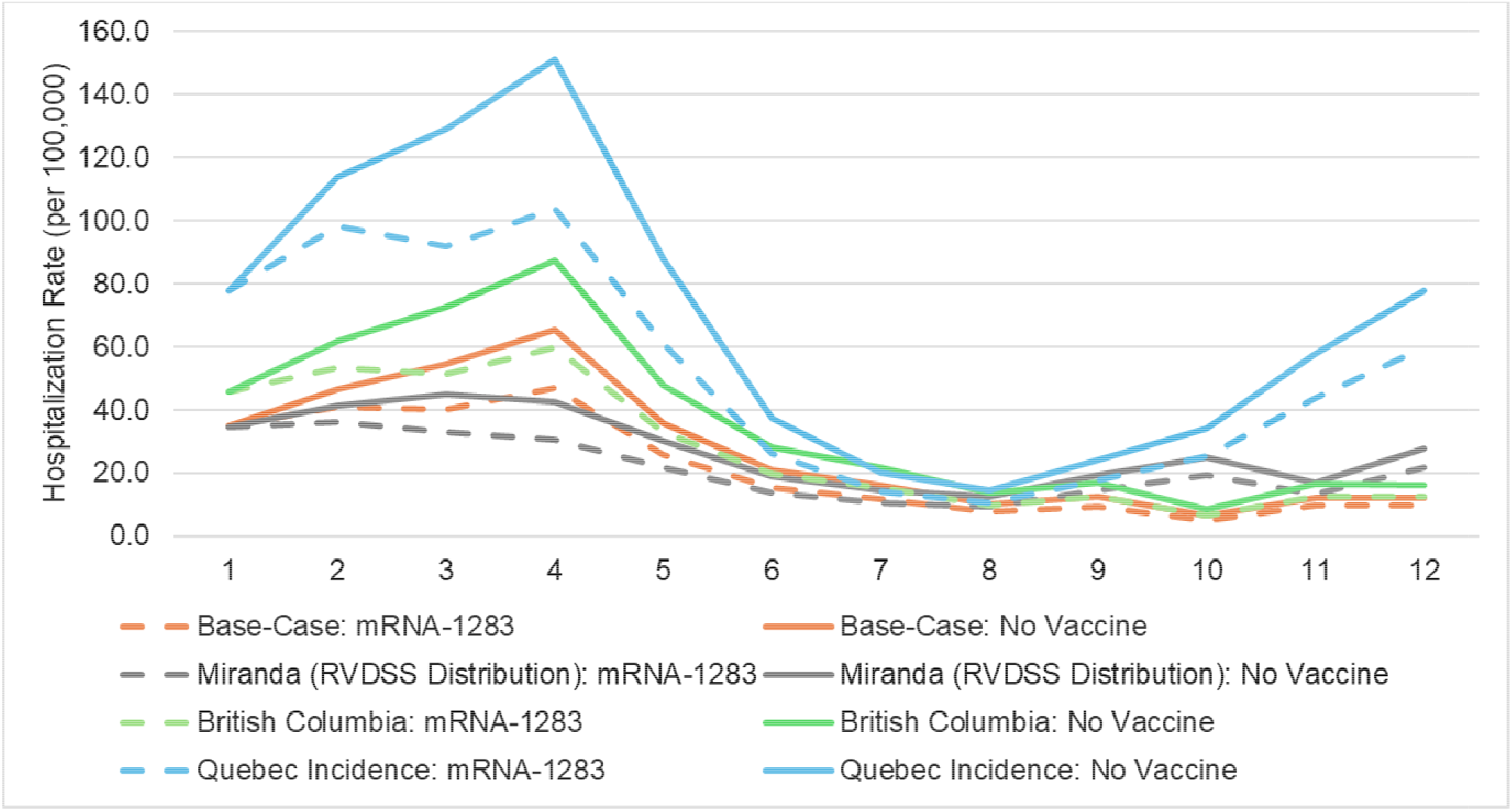
Incidence Impact on Hospitalization Rates per 100,000 (with and without vaccination)

**Figure 5.**
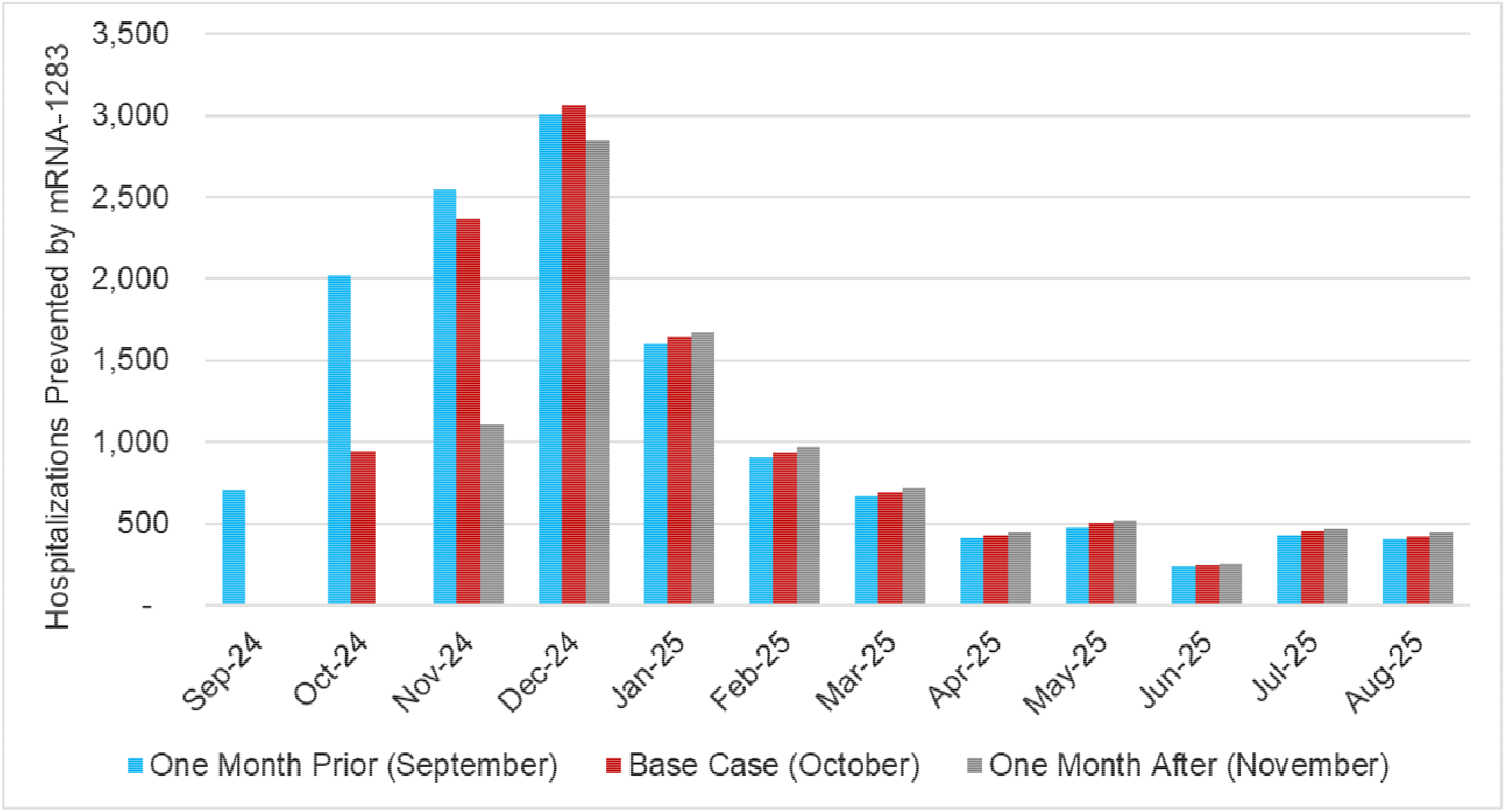
Impact of Shifting Vaccination Coverage to Start One Month Earlier (in September) and One Month Later (November) compared to the Base-Case (October) on Hospitalizations Prevented by mRNA-1283. *Apr: April; Aug: August; Dec: December; Feb: February; Jan: January; Jul: July; Jun: June; Mar: March; Nov: November; Oct: October; Sep: September*

Relative to mRNA-1273 and BNT162b2, respectively, mRNA-1283 is projected to avert an additional 2,873 and 3,689 hospitalizations, and 537 and 690 deaths (Table 4). The EJPs in these comparisons represent the premium above the cost (at any price) of the authorized vaccines at a WTP threshold of $50,000/QALY gained: $78 over mRNA-1273 and $103 over BNT162b2.

**Table 4.** Results: mRNA-1283 Compared to Existing COVID-19 Vaccines.

| Target Population | Vaccination Strategy | Outcomes |  |  |  | Outcomes Averted |  |  |  |
| --- | --- | --- | --- | --- | --- | --- | --- | --- | --- |
|  |  | Infections | Hosp. | PCC | Deaths | Infections | Hosp. | PCC | Deaths |
| *Base-case (65+ all; 12-64 HR; annual dosing)* | mRNA-1273 | 2,997,082 | 45,077 | 40,325 | 8,088 | -- | -- | -- | -- |
|  | mRNA-1283 | 2,955,676 | 42,204 | 39,521 | 7,551 | 41,407 | 2,873 | 805 | 537 |
| *Base-case (65+ all; 12-64 HR; annual dosing)* | BNT162b2 | 3,026,504 | 45,893 | 41,007 | 8,241 | -- | -- | -- | -- |
|  | mRNA-1283 | 2,955,676 | 42,204 | 39,521 | 7,551 | 70,828 | 3,689 | 1,486 | 690 |

HR, High-risk; Hosp.: Hospitalizations; PCC: Post-COVID condition (long COVID)
65+ all; 12-64 HR: Includes the population aged 12-64 years of age at high-risk, and the general population ≥65 years of age.

## Interpretation

Model projections suggest that mRNA-1283 has the potential to meaningfully reduce the clinical burden of COVID-19 for adults ≥65 years and high-risk 12-to-64-year-olds. Expanding the vaccination strategy to include semi-annual dosing for those ≥65 years or ≥80 years is expected to avert additional hospitalizations and deaths compared to annual vaccination. The EJP at a $50,000/QALY WTP threshold for the base-case analysis for mRNA-1283 compared to no vaccination is $325 ($230-$771 in scenario analyses). Results of pair-wise comparisons between mRNA-1283 and existing COVID-19 vaccines suggest that mRNA-1283 could prevent additional hospitalizations and deaths. EJPs for mRNA-1283 were $78 and $103 when compared to mRNA-1273 and BNT162b2, respectively. These results quantify the impact of differential VE between the next-generation mRNA-1283 vaccine compared to currently authorized COVID-19 vaccines and may help inform vaccine choice.

Model results were highly sensitive to the incidence of symptomatic COVID-19 infection. As public funding decisions for COVID-19 vaccines shift to provinces and territories, differences in surveillance practices, testing patterns, and in-hospital definitions of COVID-19 cases across jurisdictions may impact the evaluation of COVID-19 vaccines.(33) Model results were also sensitive to shifts in vaccine timing; shifting vaccine administration with mRNA-1283 to September would prevent an additional 34,100 symptomatic infections, 1,700 hospitalizations and 318 deaths compared to the base-case. Consistent with Miranda et al. (2024)(17), this result suggests that administering vaccination prior to peak COVID-19 incidence has a substantial benefit on the clinical impact of COVID-19 vaccines; as the seasonality of COVID-19 has not yet been fully established, vaccination timing is critical to maximize clinical benefit.

Miranda et al. (2024)(17) published a CUA examining COVID-19 vaccination strategies in Canada; several scenarios (Technical Appendix) were performed to compare the current study results to Miranda et al. The models differ in terms of approach; Miranda et al. performed a microsimulation with a focus on medically-attended cases, while our analysis utilizes a cohort model and considers those receiving no formal care. One key difference between models is VE; the current model assumes a linear decline in VE, whereas the Miranda et al. publication assumes VE declines in a step-wise fashion. VEs against medically-attended and inpatient cases used to inform the Miranda et al. model were obtained from the US Centers for Disease Control and Prevention (CDC) Advisory Committee on Immunization Practices (ACIP) presentation on the effectiveness of COVID-19 (2023-2024 formula) vaccines(27). In the strategy similar to the present study’s base-case (all aged 65+ years and high risk ≤65 years), the initial VE against emergency department/urgent care encounters and hospitalization from the VISION Network for those aged 18+ years was applied to the 65+ years group (50% and 49%, respectively). VEs from Miranda et al.(17) were based on XBB.1.5 formulas and did not differentiate between mRNA-1273 and BNT-162b2. VEs for the high-risk age groups were assumed to be 10% lower than non-high-risk.

mRNA-1273 VE estimates used in the current study projection period were product specific and based on the 2024-2025 KP.2 mRNA-1273 vaccine.(25, 26) Values used for the high-risk aged groups were based on real-world data on high-risk individuals.(29,34,35) mRNA-1283 VE estimates were based on comparative data to mRNA-1273.(35) The difference in our VE estimates for those aged 65+ compared to those used by Miranda et al. (54.5% for infection; 57.3% for hospitalization) can be explained due to a different season and variant adapted vaccine (XBB.1.5 vs KP.2) as well as being product-specific. The mRNA-1273 vaccine has been shown to result in higher VE values compared to BNT-162b2.(36-38) Furthermore, a recent Danish observational study suggests high levels of sustained protection over four months against hospitalization and death.(39) Similarly, results from the VISION network(40) suggest no waning of VE against hospitalization for the 2024/2025 mRNA vaccines in adults ≥65 years over the first four months after receipt. Finally, due to the cohort model design, the current study utilizes average VE per month, whereas in a microsimulation, the VE changes for each individual.

This study is subject to several limitations. The model uses a static approach and does not take into account indirect effects. Transmission studies have reported indirect effects(41-43), suggesting that future research addressing vaccination benefits in younger age groups should include potential indirect benefits of vaccination. While the modelling data are promising, mRNA-1283 safety and effectiveness have not yet been validated in real-world studies, and rVE estimates for mRNA-1283 compared to mRNA-1273 and BNT162b2 need to be confirmed by head-to-head comparative effectiveness studies in the real-world setting. Further, there is high uncertainty around VE waning estimates. Incidence estimates are also highly uncertain, not only due to the aforementioned differences across provinces in surveillance practices, but also as COVID-19 continues to evolve. In the absence of Canadian data, estimates from the United Kingdom and U.S. were used as proxies.

Based on this study, mRNA-1283 could potentially reduce the clinical burden of COVID-19 in the NACI-recommended population (all aged ≥65 and high-risk 12-64), offering added benefit over currently authorized COVID-19 mRNA vaccines in Canada. The additional clinical burden prevented by mRNA-1283 yields meaningful incremental health economic value at Canada’s commonly used WTP threshold(12-16) of $50,000/QALY, supporting evidence-informed vaccine selection and optimization of provincial and territorial COVID-19 vaccination programs to better protect those with the highest unmet need.

## Supporting information

Technical Appendix

## Data Availability

All data produced in the present work are contained in the manuscript

