## Supplementary material for "The Potential Clinical and Economic Impact of the Next-Generation COVID-19 mRNA-1283 Vaccine in Canada": Technical Appendix

### Infection Incidence

A simplified version of the static model is used to estimate the incidence of SARS-CoV-2 infection in those not vaccinated using the steps outlined below.

**Step 1**: Develop the age-specific targets for the monthly rate of hospitalizations by age group for a one-year period. For the base case, the hospitalization rates presented by Miranda et al. (2024) were used, along with the monthly distribution of COVID-19 cases based on the Government of Canada data. These monthly hospitalization rate targets were further adjusted to reflect the proportion of patients hospitalized with confirmed COVID with ≥1 comorbid condition as reported by Miranda et al. (2024) to estimate hospitalization rate targets specific to the high-risk population for ages <65; hospitalization rate targets for ages 65+ were not adjusted, as all adults (not just those at high-risk) are included in the analysis .

The base-case target hospitalization rates per 100,000 are displayed in Table 1 below.

Table 1. Target hospitalization rates per 100,000 for the calculation of infection incidence

| **Age Group** | **Sept** | **Oct** | **Nov** | **Dec** | **Jan** | **Feb** | **Mar** | **Apr** | **May** | **Jun** | **July** | **Aug** |
| --- | --- | --- | --- | --- | --- | --- | --- | --- | --- | --- | --- | --- |
| 12-17 | 0.78 | 0.95 | 1.00 | 1.18 | 0.65 | 0.38 | 0.29 | 0.19 | 0.23 | 0.12 | 0.23 | 0.23 |
| 18-49 | 2.12 | 2.57 | 2.71 | 3.22 | 1.76 | 1.03 | 0.80 | 0.51 | 0.63 | 0.32 | 0.63 | 0.63 |
| 50-64 | 7.78 | 9.47 | 9.96 | 11.8 | 6.48 | 3.80 | 2.93 | 1.87 | 2.30 | 1.18 | 2.30 | 2.30 |
| ≥ 65 | 63.4 | 77.1 | 81.1 | 96.3 | 52.7 | 30.9 | 23.8 | 15.2 | 18.8 | 9.63 | 18.8 | 18.8 |

**Step 2:** Enter the age-specific probability of hospitalization and the related proportion of symptomatic cases that are seeking care.

**Step 3**: Enter the assumed vaccination coverage.

The monthly vaccine coverage rate for September 2023 to August 2024, displayed in Table 2, were estimated from the vaccine coverage of the XBB.1.5 vaccines from the Government of Canada(1). Figure 1 provides detailed vaccine coverage over time for the first dose.

Figure 1. First dose COVID-19 vaccine coverage during season 2023-2024, by age groups

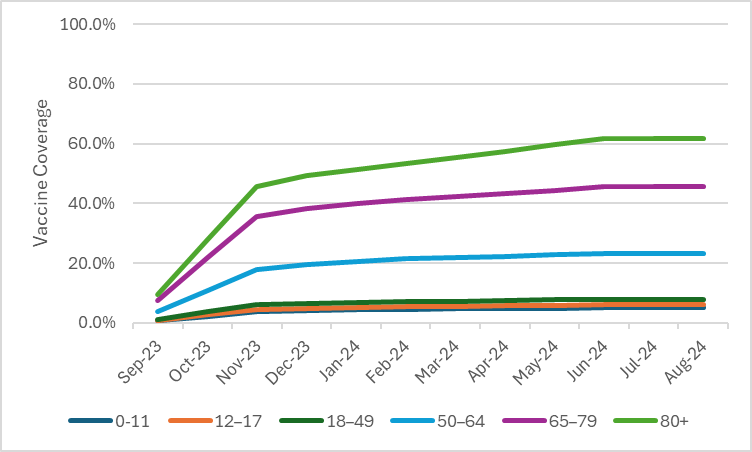

**Step 4**: Enter the assumed initial vaccine effectiveness (VE) against infection and hospitalization and the monthly linear waning rate over time.

2023-2024 COVID-19 VEs used during the calibration period were based on a market mix of the COVID-19 mRNA vaccines mRNA-1273 and BNT162b2 administered during 2023-2024. For those 18 years and over, both VE outcomes of the Moderna XBB.1.5 vaccine were estimated based on a study by Kopel et al. (2024).(2) They estimated the VE in those ≥18 years old receiving the vaccine between September 13, 2023 and December 15, 2023 relative to individuals that did not receive the XBB.1.5 vaccine during the same time period, independent of prior vaccination history. As the median follow-up time was 70 days post-vaccination, values for both outcomes were scaled back by 56 days to 14 days post vaccination to approximate the maximum level of protection assuming a monthly waning rate of 4.75% for infection (3) and 2.46% for hospitalization(4).

The 2023-2024 VE for the Pfizer XBB.1.5 vaccine were estimated based on adjusted rVE values between Moderna and Pfizer vaccine values. At time of calculation, values were available for bivalent original and Omicron BA.4/5 containing mRNA-1273.222 versus BNT1162b2. rVE for outpatient visits, considered a proxy for symptomatic infection, (rVE=5.1%, 95% CI 3.2-6.9%) and rVE for hospitalization (rVE=9.8%, 95% CI 2.6-16.4%) were used.(5) The Moderna and Pfizer 2023-2024 COVID-19 VE values were then weighed by the estimated market shares of each (46% and 54%, respectively(6)), to obtain a weighted 2023-2024 COVID-19 VE for the market mix.

For the age group 0 to 17 years, the 2023-2024 COVID-19 VE value was obtained from a study by Link-Gelles.(7) This source was also used to inform the VE for those aged 12-17 on mRNA-1273 or BNT-162b2 in the projection period. The 2023-2024 COVID-19 VE for the age group of 5-17 years was 71.0% and assumed for those 0-17 years in the calibration period. As VE was measured between 7-59 days post XBB.1.5 vaccine administration, adjustments to 2 weeks post-administration was not required.

Table 2 summarizes the mRNA-1273, BNT162b2, and market mix VE used for calculation of the incidence of symptomatic infection without vaccination.

Table 2. Market mix VE used in the calculation of infection incidence

| **Vaccine** | **Age Group** | **Infection** | | **Hospitalization** | |
| --- | --- | --- | --- | --- | --- |
|  |  | **Initial VE** | **Waning** | **Initial VE** | **Waning** |
| mRNA-1273 | 18+ years | 41.8% | 4.75% | 64.7% | 2.46% |
| BNT162b2 | 18+ years | 38.7% | 4.75% | 60.9% | 2.46% |
| Market Mix | 0 to 17 years | 71.0% | 4.75% | 71.0% | 2.46% |
| Market Mix | 18+ years | 40.1% | 4.75% | 62.7% | 2.46% |

VE: Vaccine effectiveness

**Step 5: Estimate the rates using the model decision tree calculations**

An initial set of monthly incidence rates of symptomatic infection without seasonal vaccination is used to calculate the number of hospitalizations by month and age group. These estimated hospitalization counts are compared to the target number of hospitalizations by month and age group and the ratio of target to model estimated hospitalizations is calculated for each month and age group. These ratios are multiplied by the initial set of monthly incidence rates to determine the calibrated monthly incidence rates by age group that are required to estimate the target number of hospitalizations, given the other model inputs are held constant.

The base case incidence values are displayed in Figure 2. Several scenario analyses were conducted to account for the variation of future COVID-19 rates, including:

- an alternative scenario (incidence 2) using the same annual hospitalization incidence from Miranda et al. (2024)(8), adjusted for risk status, with the monthly distribution based on the Respiratory Virus Detection Surveillance System (RVDSS) data;
- A British Columbia (BC)(9) incidence scenario was developed using annual hospitalization data from the 2023-2024 season and the Miranda et al. monthly distribution of cases; and
- a Quebec(10) incidence scenario developed using monthly hospitalization rate data from 2023-2024.

To allow for a full range in incidence variation, the BC and Quebec estimates were not adjusted for those at high-risk. Graphs displaying the various incidence scenarios are presented below.

Figure 2. Base-Case Incidence of Symptomatic COVID-19 Infection (no vaccine arm)

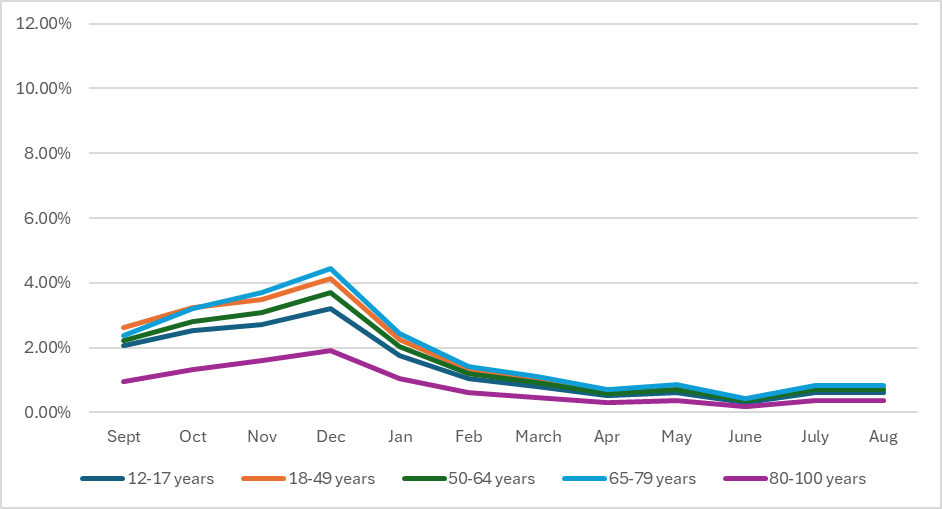

Figure 3. Scenario Analysis: Incidence of Symptomatic COVID-19 Infection based on Miranda et al. (2024) (Respiratory Virus Detection Surveillance System Distribution) (no vaccine arm)

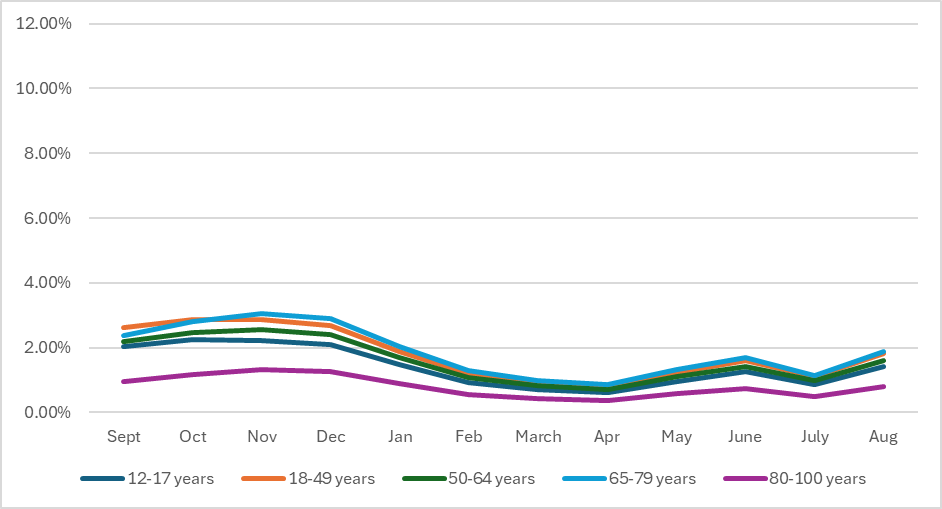

Figure 4. Scenario Analysis: Incidence of Symptomatic COVID-19 Infection based on British Columbia (no vaccine arm)

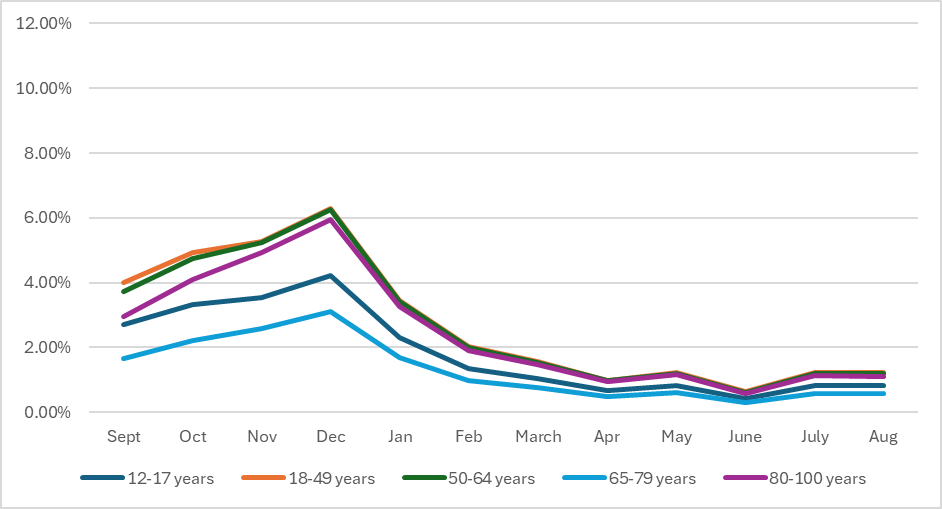

Figure 5. Scenario Analysis: Incidence of Symptomatic COVID-19 Infection based on Quebec (no vaccine arm)

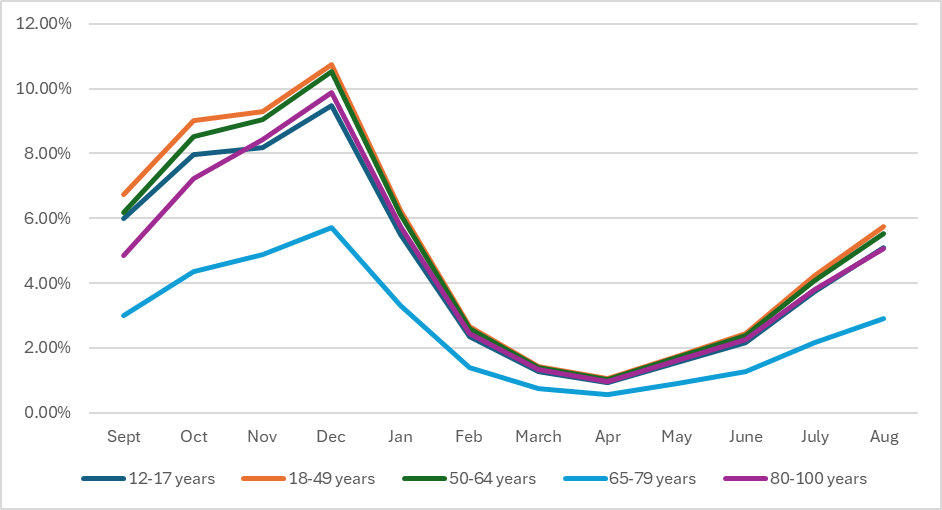

### Overview of Vaccine Effectiveness Inputs

The 2024-2025 COVID-19 VE is defined as the measure of the initial VE in individuals that received a vaccine in 2024-2025 compared to individuals that did not receive a 2024-2025 vaccine, regardless of prior vaccination history. Those that did not receive the vaccine may still have existing protection against infection and hospitalization due to prior vaccination and/or infections (i.e., the baseline level of protection in the population before receiving the new vaccine).

To project the impact of the 2024-2025 mRNA-1283 vaccine, the annual VE against infections and hospitalizations is required. To estimate the initial VEs of mRNA-1283, age-specific rVEs of mRNA-1283 compared to mRNA-1273 was applied to the 2024-2025 VEs of mRNA-1273.

2024-2025 VEs of mRNA-1273 against infection and hospitalization for ages 18-64 years were based on high-risk individuals aged 18+ years from a study by Wilson et al., (2025). VEs for those aged 65+ years were based on all individuals aged 65+ years from the same study. This study estimated the VE against medically-attended COVID-19 (used as a proxy for infections) and COVID-19 related hospitalization in adults that received a dose of mRNA-1273.712 targeting KP.2 during the 2024-2025 season compared to those that did not receive any COVID-19 vaccine during the 2024-2025 season vaccine.(11, 12) Because VEs were measured at 64 days post administration, the values were scaled back to 14 days post administration, to represent the starting, or highest VE achieved after administration. This adjustment was done assuming a monthly VE waning rate of 4.75% against infection(3) and 2.46% against hospitalization.(4)

For those aged 12-17 years, due to the lack of differentiating VE data for infections and hospitalizations, and for data on the KP.2 variant, the initial 2024-2025 VEs for mRNA-1273 against infection and hospitalization were assumed to be the same and were based on estimates by Link-Gelles et al. (2024) of the VE of XBB.1.5 vaccines against emergency department and urgent care visits in children aged 5-17 years,(7).

Comparative VE data (relative VE,rVE) values for mRNA-1283 compared to mRNA-1273 (rVE = 17.5% for infection in high-risk individuals aged 12-64 years, 13.65% in those age 65+ years, and 38.06% for hospitalizations) were applied to the 2024-2025 mRNA-1273 VEs against KP.2 to calculate the annual mRNA-1283 VE values.(13) These values were obtained from the NextCOVE phase 3 pivotal study (Study P301). rVE data for hospitalizations in high-risk groups were not available and therefore the value for all individuals aged 12+ years was used. Note that the rVE against COVID-19 CDC defined symptomatic infections(14) was used as proxy for the rVE against infection. For those aged 80+ years, values for the ≥65+ years age group are used for the second dose.

To address the second objective, initial 2024-2025 BNT-162b2 VEs against infection and hospitalization were needed. VEs assumed to be the same as the 2024-2025 mRNA-1273 values for those age 12-17 years, as the published values(7) were based on a mix of mRNA-1273 and BNT-162b2. BNT-162b2 VE values against infection for those aged 18-64 years were calculated using the rVE of mRNA-1283 compared to BNT-162b2 for high-risk individuals aged 18+ years (20.53%) from an indirect treatment comparison by Beck et al. (2025).(15, 16) The rVE for those aged 65+ (22.80%) was also obtained from the same source, but for all those aged 65+. The rVE value against hospitalization was not available for high-risk individuals and therefore the rVE for all individuals aged 12+ years (44.13%) was applied for all ages.

Deterministic sensitivity analyses were conducted on initial VE. In the case of the comparison between mRNA-1283 and no seasonal vaccination, this included using the upper and lower 95% confidence intervals (CIs) of mRNA-1283 as the initial VE, applying the same rVE for mRNA-1283/mRNA-1273 for infection to hospitalization. The upper and lower 95% CIs for the VE of mRNA-1283 against infection and hospitalization were derived based on Monte-Carlo simulations considering the 95% CIs of the VE of mRNA-1273 and the rVE estimates. Finally, a sensitivity analysis was included using the same starting VEs used by Miranda et al. (2024)(8) in their recently published Canadian cost-effectiveness analysis of COVID-19 vaccines. These VE values were obtained from the US Centers for Disease Control and Prevention (CDC) Advisory Committee on Immunization Practices (ACIP) presentation on the effectiveness of COVID-19 (2023-2024 formula) vaccines.(7)

### Equations for Incremental Vaccines Effectiveness

For each age group, we define the following vaccine effectiveness variables and relationship between the variables. The superscripts and subscripts for age group are removed for clarity.

Definitions

${VE}_{1}$ = Vaccine effectiveness against infection

${VE}_{2}$ = ‘Total’ Vaccine effectiveness against hospitalization

${VE}_{2}^{*}$= ‘Additional’ Vaccine effectiveness against hospitalization

We assume ${VE}_{2}^{*}=0$ if there is no additional benefit against hospitalization

Define

$$\left[ 1-{VE}_{2} \right]= \left[ 1-{VE}_{1} \right]\times\left[ 1-{VE}_{2}^{*} \right]$$

Isolate and solve for ${VE}_{2}^{*}$

$$\left[ 1-{VE}_{2}^{*} \right]=\frac{\left[ 1-{VE}_{2} \right]}{\left[ 1-{VE}_{1} \right]}$$

$${VE}_{2}^{*}= 1- \frac{\left[ 1-{VE}_{2} \right]}{\left[ 1-{VE}_{1} \right]}$$

### Calculation of Economically Justifiable Price (EJP)

Economically justifiable price (EJP) is calculated through manipulation of the standard formula for the incremental cost-effectiveness ratio (ICER). The cost of the 1283 vaccine is calculated so that the ICER within the model will equal the willingness to pay (WTP) threshold. For this analysis, the costs of administration and wastage were included in the total costs meaning that the EJP represents the unit cost per dose of vaccine.

$$WTP=willingness to pay$$

$$ICER=incremental cost-effectiveness ratio$$

$$\Delta QALYs=-1 \times( {QALYs Lost}_{1283}-{QALYs Lost}_{No Vaccine})$$

$$\Delta Costs={Costs}_{1283}-{Costs}_{No Vaccine}$$

$${Costs}_{1283}={Costs}_{1283 Vaccination}+{Costs}_{1283 Disease}$$

$${EJP}_{1283 vaccination}={EJP}_{1283 vaccine}+{Costs}_{1283 administration}$$

$$ICER= \frac{\Delta Costs}{\Delta QALYs}$$

Set the $ICER=WTP$ for calculation of $EJP$.

$$WTP= \frac{\Delta Costs}{\Delta QALYs}$$

$$\Delta Costs= WTP \times\Delta QALYs$$

$${Costs}_{1283}-{Costs}_{No Vaccine}=WTP \times\Delta QALYs$$

$${Costs}_{1283}=WTP \times\Delta QALYs+{Costs}_{No Vaccine}$$

$${Costs}_{1283 vaccination}+{Costs}_{1283 Disease}= WTP \times\Delta QALYs+{Costs}_{No Vaccine}$$

$${Costs}_{1283 vaccination}= WTP \times\Delta QALYs+{Costs}_{No Vaccine}-{Costs}_{1283 Disease}$$

This will give us the maximum costs available for the cohort for vaccination, including vaccine administration costs. The individual vaccination cost is calculated based on the number of vaccinations estimated in the model for 1283.

$${EJP}_{1283 vaccination}=\frac{WTP \times\Delta QALYs+{Costs}_{No Vaccine}-{Costs}_{1283 Disease}}{Number of Vaccinations}$$

$${EJP}_{1283 vaccine}=\frac{WTP \times\Delta QALYs+{Costs}_{No Vaccine}-{Costs}_{1283 Disease}}{Number of Vaccinations}-{Costs}_{1283 administration}$$

### Model Inputs

Model inputs are presented in the tables below.

Table 3. Model Inputs: Target Population Size

| **Age Group** | **Canadian Population** | **Percentage Eligible*** | **Target Population Size** | **References** |
| --- | --- | --- | --- | --- |
| 12-17 years | 2,702,398 | 21.0% | 567,504 | Statistics Canada(17); Queenan 2021(18)  Statistics Canada(19) |
| 18-49 years | 18,191,799 | 24.9% | 4,529,758 | Statistics Canada(17); Queenan 2021(18)  Statistics Canada(19) |
| 50-64 years | 7,653,083 | 45.6% | 3,489,806 | Statistics Canada(17); Queenan 2021(18)  Statistics Canada(19) |
| 65-79 years | 5,948,066 | 100% | 5,948,066 | Statistics Canada(17) |
| 80+ years | 1,872,055 | 100% | 1,872,055 | Statistics Canada(17) |

*****Percentage eligible for those <65 years reflects proportion at high-risk for severe outcomes from COVID-19; estimates are consistent with Miranda et al. (2024) and reflect the proportion of the population with one or more chronic medical conditions

Table 4. Model Inputs: Vaccine Coverage (Projection Period)

| **Month** | **12-17 years** | **18-49 years** | **50-64 years** | **65-79 years** | **80+ years** | **References** |
| --- | --- | --- | --- | --- | --- | --- |
| September | 0.0% | 0.0% | 0.0% | 0.0% | 0.0% | National Institute of Public Health of Quebec(20) |
| October | 0.4% | 1.7% | 5.1% | 16.2% | 23.0% |  |
| November | 1.9% | 5.0% | 12.5% | 35.2% | 49.2% |  |
| December | 2.4% | 5.9% | 14.3% | 39.1% | 54.3% |  |
| January | 2.5% | 6.1% | 14.6% | 39.8% | 55.1% |  |
| February | 2.6% | 6.2% | 14.8% | 40.2% | 55.4% |  |
| March | 2.6% | 6.2% | 14.9% | 40.4% | 55.7% |  |
| April* | 2.6% | 6.2% | 14.9% | 40.4% | 55.7% |  |

*Estimates assumed to apply from April forward (through August)

Table 5. Model Inputs: Vaccine Coverage for Second Dose (Projection Period)

| **Month** | **12-17 years** | **18-49 years** | **50-64 years** | **65-79 years** | **80+ years** | **References** |
| --- | --- | --- | --- | --- | --- | --- |
| February | 0.0% | 0.0% | 0.0% | 0.0% | 0.0% | National Institute of Public Health of Quebec(20): Miranda et al. (2024)(8) |
| March | 0.0% | 0.0% | 0.0% | 3.39% | 4.81% |  |
| April | 0.0% | 0.0% | 0.0% | 7.36% | 10.28% |  |
| May | 0.0% | 0.0% | 0.0% | 8.17% | 11.35% |  |
| June | 0.0% | 0.0% | 0.0% | 8.45% | 11.64% |  |
| July | 0.0% | 0.0% | 0.0% | 8.45% | 11.64% |  |
| August | 0.0% | 0.0% | 0.0% | 8.45% | 11.64% |  |

Table 6. Model Inputs: Probabilities

| **Parameter** | **Base** | **Range** | **Reference** | |
| --- | --- | --- | --- | --- |
| **Percentage hospitalized, given infection*** | | | | |
| **12-17 years (high-risk)** | 0.06% | ±10% | | Derived based on UK data(21); Adjusted to reflect the unvaccinated population |
| **18-49 years (high-risk)** | 0.12% |  |  |  |
| **50-64 years (high-risk)** | 0.46% |  |  |  |
| **65-79 years (all)** | 2.79% |  |  |  |
| **80+ years (all)** | 7.04% |  |  |  |
| **Percentage with no formal care, given infection*** | | | | |
| **12-17 years (high-risk)** | 94.33% | -- | | Based on US CDC influenza data; calculated as 1 - percentage of hospitalization and outpatient care for high-risk patients |
| **18-49 years (high-risk)** | 94.35% | -- | |  |
| **50-64 years (high-risk)** | 89.64% | -- | |  |
| **65-79 years (all)** | 74.68% | -- | |  |
| **80+ years (all)** | 38.50% | -- | |  |
| **In-hospital distribution across locations of care (%) (all ages)** | | | | |
| **No ICU or Ventilator** | Calculated |  | |  |
| **ICU only** | 8.1% | 7.7%-8.4% | | CIHI(22) |
| **ICU with Ventilator** | 5.1% | 4.8%-5.4% | | CIHI(22) |
| **In-hospital mortality (%) (No ICU or ventilator, ICU only, ICU with ventilator)**^†^ | | | | |
| **12-17 years (high-risk)** | 2.9% | 2.18%-3.63% | | Miranda 2024(8) |
| **18-49 years (high-risk)** | 3.3% | 2.48%-4.13% | |  |
| **50-64 years (high-risk)** | 9.5% | 7.13%-11.88% | |  |
| **65-79 years (all)** | 14.8% | 11.09%-18.48% | |  |
| **80+ years (all)** | 17.9% | 13.43%-22.38% | |  |
| **Hospital readmission (%) (No ICU or ventilator, ICU only, ICU with ventilator)** | | | | |
| **All ages** | 0.0% | -- | | Assumed to be 0% to avoid double-counting with hospitalization cost estimates |
| **Post-discharge mortality (%) (No ICU or ventilator, ICU only, ICU with ventilator)** | | | | |
| **All ages** | 3.7% | 3.5%-3.9% | | Qian 2025(23) |
| **Infection-Related Myocarditis** | | | | |
| **% Female** |  |  | |  |
| **12-17 years (high-risk)** | 50% | -- | | Assumption |
| **18-49 years (high-risk)** | 50% |  |  |  |
| **50-64 years (high-risk)** | 50% |  |  |  |
| **65-79 years (all)** | 50% |  |  |  |
| **80+ years (all)** | 50% |  |  |  |
| **% Infection-Related Myocarditis** | | | | |
| **Female** |  |  | |  |
| **12-17 years (high-risk)** | 0.122% | -- | | Boehmer 2021(24) |
| **18-49 years (high-risk)** | 0.079% |  |  |  |
| **50-64 years (high-risk)** | 0.137% |  |  |  |
| **65-79 years (all)** | 0.172% |  |  |  |
| **80+ years (all)** | 0.208% |  |  |  |
| **Male** |  |  | |  |
| **12-17 years (high-risk)** | 0.122% | -- | | Boehmer 2021(24) |
| **18-49 years (high-risk)** | 0.079% |  |  |  |
| **50-64 years (high-risk)** | 0.137% |  |  |  |
| **65-79 years (all)** | 0.172% |  |  |  |
| **80+ years (all)** | 0.208% |  |  |  |
| **Post-Acute Care** |  |  | |  |
| **Hospitalized or Received Outpatient Care** | | | | |
| **% Post-Infection** |  |  | |  |
| **12-17 years (high-risk)** | 100% | -- | | Assumption(consistent with post-infection QALY loss estimates) |
| **18-49 years (high-risk)** | 100% |  |  |  |
| **50-64 years (high-risk)** | 100% |  |  |  |
| **65-79 years (all)** | 100% |  |  |  |
| **80+ years (all)** | 100% |  |  |  |
| **% Long COVID^‡^** |  |  | |  |
| **12-17 years (high-risk)** | -- | -- | | CDC 2025(25) |
| **18-49 years (high-risk)** | 8.5% | 7.0%-10.2% | |  |
| **50-64 years (high-risk)** | 9.6% | 8.4%-11.0% | |  |
| **65-79 years (all)** | 7.3% | 6.2%-8.6% | |  |
| **80+ years (all)** | 9.6% | 5.2%-15.7% | |  |
| **% Severe Long COVID** |  |  | |  |
| **12-17 years (high-risk)** | 0% | -- | | Assumption |
| **18-49 years (high-risk)** | 0% |  |  |  |
| **50-64 years (high-risk)** | 0% |  |  |  |
| **65-79 years (all)** | 0% |  |  |  |
| **80+ years (all)** | 0% |  |  |  |
| **Canadian Life Tables** |  |  | |  |
| **All ages** | Values are age-dependent | -- | | Statistics Canada(26) |

ICU: Intensive care unit

*Estimates for high-risk patients derived using a RR=1.69 for hospitalization and RR=1.17 for outpatient care based on Moderna claims and EHR data (Veradigm database) retrospective observational analysis

^†^Death is assumed to occur via hospitalization only

^‡^Applies to analyses performed from the societal perspective only

Table 7. Model Inputs: Healthcare Costs

| **Parameter** | **Base** | **Range** | **Reference** |
| --- | --- | --- | --- |
| **Vaccine administration cost** | | | |
| **Wastage** | 10% | -- | Assumption based on Miranda 2025 |
| **Cost per administration** | $11.40 | -- | OHIP 2025(27) |
| **Adverse Event Costs** |  |  |  |
| **Grade 3 Local or Systemic** | $0.77 | -- | NextCOVE  Ontario Ministry of Health(27-29)  Rousculp 2024(30)  Walmart 2025(31)  CIHI 2021(32) |
| **Grade 4 Local** | -- | -- |  |
| **Grade 4 Systemic** | $64 | -- | Calculated based on Miranda 2024(8) |
| **Anaphylaxis** | $15,079 | -- | CIHI(33) |
| **Myocarditis/Pericarditis** | $10,293 | -- | CIHI(33) |
| **Infection-Related myocarditis cost** | | | |
| **Infection-related myocarditis event** | $10,293 | -- | CIHI(33) |
| **Infection-Related Costs** |  |  |  |
| **No Formal Health Care** | $0 |  |  |
| **Outpatient Care** | $302 | $212-$392 | Sander 2025(34) |
| **Initial Hospitalization Costs*** | | | |
| **No ICU or Ventilator** | $30,876 | $29,456-$32,297 | Sander 2025(34) |
| **ICU only** | $108,233 | $102,897-$113,570 | Sander 2025(34) |
| **ICU with Ventilator** | $108,233 | $102,897-$113,570 | Sander 2025(34) |

ICU: Intensive care unit

*Costs reflect a 1-year follow-up period based on Sander et al. (2025); accordingly, costs of hospital readmission, recovery, and post-COVID conditions (including Long COVID) are assumed to be reflected in the initial hospitalization costs

Table 8. Model Inputs: Patient Productivity Losses (Societal Perspective Only)

| **Parameter** | **Base** | **Range** | **References** |
| --- | --- | --- | --- |
| **Percentage in Labor Force** | | | |
| **12-17 years (high-risk)** | 19.5% | -- | Statistics Canada(35) |
| **18-49 years (high-risk)** | 79.3% | -- |  |
| **50-64 years (high-risk)** | 70.3% | -- |  |
| **65-79 years (all)** | 16.2% | -- |  |
| **80+ years (all)** | 8.0% | -- |  |
| **Daily wage** | | | |
| **All ages** | $252 | -- | Statistics Canada(36) |
| **Time Loss (days) due to:** | | | |
| **Vaccination** | 0.06 | -- | Miranda et al. 2024(8) |
| **Adverse Events:** |  |  |  |
| **Grade 3 Local** | 0.19 | -- | Rousculp 2024(30) |
| **Grade 4 Local** | -- | -- |  |
| **Grade 3 Systemic** | 0.19 | -- | Rousculp 2024(30) |
| **Grade 4 Systemic** | -- | -- |  |
| **Anaphylaxis** | 5.86 | -- | CIHI(22) |
| **Myocarditis/pericarditis** | 3.67 | -- | CIHI(22) |
| **Infection-related myocarditis event** | 3.67 | -- | CIHI(22) |
| **Time Loss (days): Short-term infection period** | | | |
| **No formal care** | 0 | -- | Assumption |
| **Outpatient care** | 3.57 | -- | Miranda et al. 2024(8) |
| **Hospitalization*** |  |  |  |
| **No ICU or ventilator** | 18.57 | -- | CIHI(22) |
| **ICU only** | 25.29 | -- |  |
| **ICU with Ventilator** | 25.29 | -- |  |
| **Post-acute care** |  |  |  |
| **Post-Infection** |  |  |  |
| **No Formal Health Care** | 0 | -- | Assumption |
| **Outpatient Care** | 0 | -- |  |
| **Hospitalized** | 0 | -- |  |
| **Long COVID** |  |  |  |
| **No Formal Health Care** | 0 | -- | Assumption |
| **Outpatient Care** | 23.3 | -- | Miranda 2024(8) |
| **Hospitalized** | 23.3 | -- | Miranda 2024(8) |
| **Severe Long COVID** |  |  |  |
| **No Formal Health Care** | 0 | -- | Assumption (to avoid double-counting with Long COVID) |
| **Outpatient Care** | 0 | -- |  |
| **Hospitalized** | 0 | -- |  |

ICU: Intensive care unit

*Assumed to include time loss associated with symptoms prior to hospitalization and hospitalization recovery; readmission time loss assumed equal to original location of care

Table 9. Model Inputs: QALYs Lost

| **Parameter** | **Base** | **Range** | **References** |
| --- | --- | --- | --- |
| **Adverse Events** |  |  |  |
| **Grade 3 Local or Systemic** | 0.0004 | -- | Walter 2024(37) |
| **Grade 4 Local** | -- | -- |  |
| **Grade 4 Systemic** | 0.0027 |  | Miranda 2024(8) |
| **Anaphylaxis** | 0.00192 | -- | Prosser 2019(38) |
| **Myocarditis / Pericarditis*** | 0.00064 | -- | Prosser 2019(38) |
| **Infection-Related Myocarditis Event** | 0.00192 | -- | Prosser 2019(38) |
| **Infection-Related** |  |  |  |
| **12-17 years of age** |  |  |  |
| **No Formal Care** | 0.0057 | 0.0030-0.0085 | UM-CDC(39) |
| **Outpatient Care Only** | 0.0057 | 0.0030-0.0085 | UM-CDC(39) |
| **Hospitalization*:** |  |  |  |
| **No ICU or Ventilator** |  |  |  |
| **In-hospital** | 0.0189 | 0.0054-0.0325 | UM-CDC(39) |
| **ICU only** |  |  |  |
| **In-hospital** | 0.0883 | 0.0632-0.1169 | UM-CDC(39) |
| **ICU with Ventilator** |  |  |  |
| **In-hospital** | 0.0883 | 0.0632-0.1169 | UM-CDC(39) |
| **18+ years of age** |  |  |  |
| **No Formal Care** | 0.0046 | 0.0018-0.0074 | UM-CDC(39) |
| **Outpatient Care Only** | 0.0046 | 0.0018-0.0074 | UM-CDC(39) |
| **Hospitalization*:** |  |  |  |
| **No ICU or Ventilator** |  |  |  |
| **In-hospital** | 0.0174 | 0.0038-0.0310 | UM-CDC(39) |
| **ICU only** |  |  |  |
| **In-hospital** | 0.0394 | 0.0231-0.0583 | UM-CDC(39) |
| **ICU with Ventilator** |  |  |  |
| **In-hospital** | 0.0394 | 0.0231-0.0583 | Assumption |
| **Post-acute care** |  |  |  |
| **Post-Infection** |  |  |  |
| **No Formal Health Care** | 0 | Post-Infection (all): 0  Long COVID:  0.061 (outpatient) and 0.071 (hospitalized) based on Miranda et al. (2024) | Assumption |
| **Outpatient Care** | 0.023 |  | Sandmann 2022(40) |
| **Hospitalized** | 0.103 |  | PHOSP 2022(41) |
| **Long COVID** |  |  |  |
| **No Formal Health Care** | 0 |  | Assumption |
| **Outpatient Care** | 0 |  | Assumption |
| **Hospitalized** | 0 |  | Assumption |
| **Severe Long COVID** |  |  |  |
| **No Formal Health Care** | 0 | -- | Assumption |
| **Outpatient Care** | 0 | -- | Assumption |
| **Hospitalized** | 0 | -- | Assumption |
| **Baseline Utility Data** |  |  |  |
| **12-17 years (high-risk)** | 0.886 | 0.8810-0.8900 | Guertin 2018(42) |
| **18-49 years (high-risk)** | 0.890 | 0.8836-0.8968 |  |
| **50-64 years (high-risk)** | 0.844 | 0.8363-0.8509 |  |
| **65-79 years (all)** | 0.827 | 0.8194-0.8344 |  |
| **80+ years (all)** | 0.692 | 0.6759-0.7078 |  |

CADTH, Canadian Agency for Drugs and Technologies in Health;
CIHI, Canadian Institute for Health Information;
CMDB, Canadian MIS (Management Information System) Database;
ICU, Intensive Care Unit;
OHIP, Ontario Health Insurance Plan

*Assumed to include symptoms prior to hospitalization and hospitalization recovery; readmission assumed to be equivalent to original location of care

Table 10. Model Inputs: Adverse Event Rates

| **Parameter** | **Base** | **Range** | **References** |
| --- | --- | --- | --- |
| **Adverse Event Rates (mRNA-1283)** | | | |
| **Grade 3 Local** | 1.61% | -- | NextCOVE trial data |
| **Grade 4 Local** | 0% | -- | NextCOVE trial data |
| **Grade 3 Systemic** | 7.16% |  | NextCOVE trial data |
| **Grade 4 Systemic** | 0% | -- | NextCOVE trial data |
| **Anaphylaxis** | 0.0005% | -- | Klein et al. (2021)(43) |
| **Myocarditis / Pericarditis**  **(18-49 years only)** | 0% | -- | NextCOVE trial data |
| **Adverse Event Rates (mRNA-1273)** | | | |
| **Grade 3 Local** | 1.17% | -- | NextCOVE trial data |
| **Grade 4 Local** | -- | -- | NextCOVE trial data |
| **Grade 3Systemic** | 5.77% |  | NextCOVE trial data |
| **Grade 4 Systemic** | 0.02% | -- | NextCOVE trial data |
| **Anaphylaxis** | 0.0005% | -- | Klein et al. (2021)(43) |
| **Myocarditis / Pericarditis (18-49 years only)** | 0.0018% | -- | NextCOVE trial data |
| **Adverse Event Rates (BNT162b2)** | | | |
| **Grade 3 Local** | 1.17% | -- | Assumed equal to mRNA-1273* |
| **Grade 4 Local** | -- | -- |  |
| **Grade 3 Systemic** | 5.77% |  |  |
| **Grade 4 Systemic** | -- | -- |  |
| **Anaphylaxis** | 0.0005% | -- |  |
| **Myocarditis / Pericarditis**  **(18-49 years only)** | 0.0018% | -- |  |

*No grade 4 systemic AE assumed for BNT162b2

Figure 6. Scenario Analysis: Shifting Vaccine Uptake to Start One Month Earlier (September) and One Month Later (November) compared to the Base-Case (October)

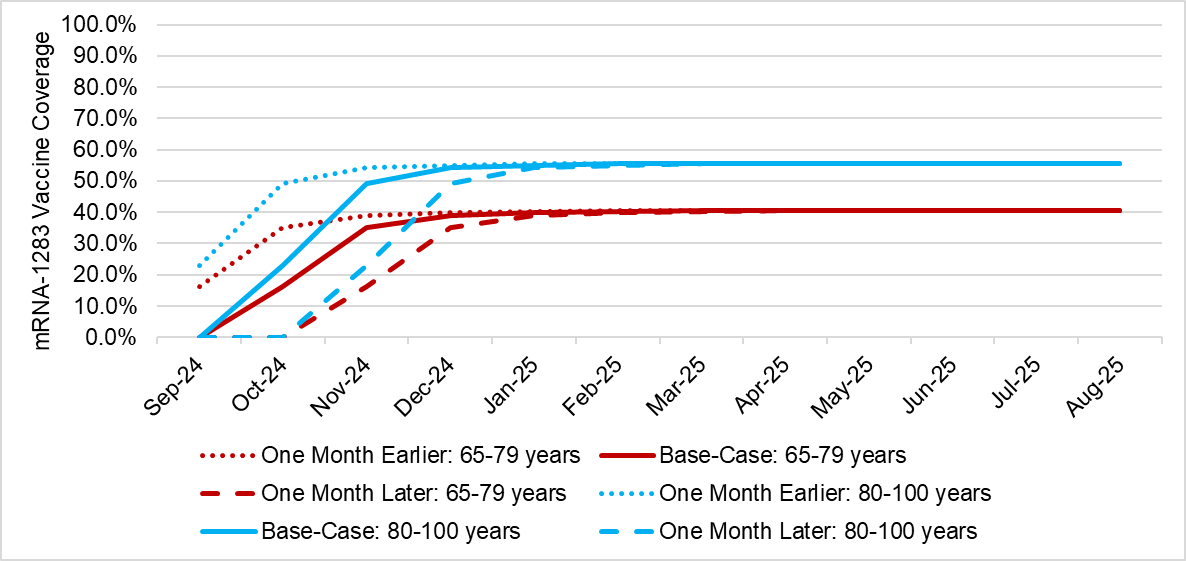

### Comparison to Miranda et al.

In order to compare our model to the previously published economic analysis conducted by Miranda et al., the model inputs from the current analysis were modified to match as closely as possible. We first compared the predictions without vaccination with both models. They were not equivalent because of the differences in model structures and some of the details required to match inputs were not available in the manuscript. Overall, the rate of hospitalizations were higher for the current model with the base-case assumptions compared to the Miranda model (average rate: 148 per 100,000 compared to 133 per 100,000). The difference was greater for outpatient care cases (1435 per 100,000 compared to 8665 per 100,000). However, the Miranda model does not track cases of symptomatic infection that do not receive formal healthcare, as the emphasis is on medically attended cases only. The current model predicted higher rates of COVID-19 attributable death: 27 per 100,000 compared to 16 per 100,000 reported by Miranda. The major difference here is the current model includes rates of increased mortality following discharge from a hospitalization attributable to COVID-19, which has been observed in multiple studies,(44) and the Miranda model does not.

As discussed in the interpretation section, the main difference between the current model and the Miranda model is the VE over time. To continue the comparisons, the current model VE was decreased to approximate the Miranda model inputs. It was also assumed that 50% of vaccines were delivered in October and 50% in November as in their analyses. Considering the base case incidence pattern, the EJP at WTP threshold of $50,000 per QALY gained was $177. When a second analyses was conducted with incidence pattern 2, the EJP decreased further to $137. All other inputs were set to the base-case estimates when these scenarios were performed.

9.	BC Centre for Disease Control. Provincial Health Services Authority. COVID-19 mRNA Vaccine Spikevax® XBB.1.5 [Available from: http://www.bccdc.ca/resource-gallery/Documents/Guidelines%20and%20Forms/Guidelines%20and%20Manuals/Epid/CD%20Manual/Chapter%202%20-%20Imms/Part4/COVID-19-mRNA-Vaccine-Spikevax-XBB-1-5.pdf.

10.	National Institute of Public Health of Quebec. COVID-19 data in Quebec 2025 [Available from: <https://www.inspq.qc.ca/covid-19/donnees>.

11.	Wilson A, Bogdanov A, Kopel H, Zheng Z, Winer I, Winer-Jones JP, et al. Interim Vaccine effectiveness mRNA-1273 KP.2 season 2024/2025. Manuscript under development. Moderna sponsored study. 2025.

12.	Wilson A BA, Zheng Z, Ryan T, Zeng N, Joshi K, Lu T, Bonafede M, Araujo AB. Evaluating the Effectiveness of 2024-2025 Seasonal mRNA-1273 Vaccination Against COVID-19-Associated Hospitalizations and Medically Attended COVID-19 among adults aged ≥18 years in the United States. medRxiv. 2025;2025.03.27.25324770.

13.	Rizkalla B. Overview of Moderna’s Investigational Next Generation COVID-19 Vaccine, mRNA-1283, in Individuals ≥12 Years of Age 2025 [Available from: https://www.cdc.gov/acip/downloads/slides-2025-04-15-16/02-Rizkalla-COVID-508.pdf.

14.	Moderna Inc. mRNA-1283 P301 Clinical Study Report.

15.	Beck E GM, Wang WJ, Gomez-Lievano A, Wang H, Gao Y, Hille B, McQueeney P, Kopel H, Bausch-Jurken M, Patterson-Lomba O, Mu F, Wu E, Van de Velde N, . An Indirect Treatment Comparison of COVID-19 Next Generation mRNA-1283 Vaccine and BNT162b2 Vaccine Against COVID-19 Symptomatic Infections in the US. AMCP; March 31 - April 3; Houston, Texas, USA2025.

16.	Beck E, Georgieva M, Wang WJ, Gomez-Lievano A, Wang H, Gao Y, et al. Indirect comparison of the relative vaccine effectiveness of mRNA-1283 vs. BNT162b2 vaccines against symptomatic COVID-19 among US adults. Curr Med Res Opin. 2025;41(4):721-32.

17.	Statistics Canada. Table 17-10-0005-01  Population estimates on July 1, by age and gender [Available from: <https://www150.statcan.gc.ca/t1/tbl1/en/tv.action?pid=1710000501>.

18.	Queenan J, Wong S, Barber D, Morkem R, Salman A. The Prevalence of Common Chronic Conditions Seen in Canadian Primary Care: Results from the Canadian Primary Care Sentinel Surveillance Network. May 2021.

19.	Statistics Canada. Table 13-10-0777-01. Number and percentage of adults (aged 18 years and older) in the household population with underlying health conditions, by age and sex (two-year period) [Available from: <https://www150.statcan.gc.ca/t1/tbl1/en/tv.action?pid=1310077801>.

20.	National Institute of Public Health of Quebec. COVID-19 vaccination data in Quebec. [Available from: <https://www.inspq.qc.ca/covid-19/donnees/vaccination>.

21.	Ward T, Fyles M, Glaser A, Paton RS, Ferguson W, Overton CE. The real-time infection hospitalisation and fatality risk across the COVID-19 pandemic in England. Nat Commun. 2024;15(1):4633.

22.	Canadian Institute for Health Information. Patient Cost Estimator [Available from: https://www.cihi.ca/en/patient-cost-estimator.

23.	Qian C, Johnston KM, Tinajero M, Voss ML, Nam A, Hamilton MA. Characterizing the clinical and economic burden of COVID-19 among individuals with immunocompromising conditions in Ontario, Canada - a matched, population-based observational study. J Med Econ. 2025;28(1):479-93.

24.	Boehmer TK, Kompaniyets L, Lavery AM, Hsu J, Ko JY, Yusuf H, et al. Association Between COVID-19 and Myocarditis Using Hospital-Based Administrative Data - United States, March 2020-January 2021. MMWR Morbidity and mortality weekly report. 2021;70(35):1228-32.

25.	Centers for Disease Control and Prevention. Post-COVID Condtions 2024 [updated October 4, 2024. Available from: <https://data.cdc.gov/NCHS/Post-COVID-Conditions/gsea-w83j/about_data>.

26.	Statistics Canada. Table 13-10-0837-01. Life expectancy and other elements of the complete life table, single-year estimates, Canada, all provinces except Prince Edward Island. [Available from: <https://www150.statcan.gc.ca/t1/tbl1/en/tv.action?pid=1310083701>.

27.	Ontario Ministry of Health. Schedule of Benefit: Physician Services. (February 14, 2025 (effective March 3, 2025)). [Available from: https://www.ontario.ca/files/2024-08/moh-schedule-benefit-2024-08-30.pdf.

28.	Ontario Drug Benefit Formulary/ Comparative Drug Index, effective from March 31, 2025. [Available from: <https://www.formulary.health.gov.on.ca/formulary/>.

29.	Ontario Ministry of Health. Health Programs and Delivery Division. Notice from the Executive Officer: Amendments to Ontario Regulation 201/96 made under the Ontario Drug Benefit Act to Change Mark-ups Paid to Dispensers. [Available from: https://www.ontario.ca/files/2024-03/moh-executive-officer-notice-change-mark-ups-en-2024-03-28.pdf.

30.	Rousculp MD, Hollis K, Ziemiecki R, Odom D, Marchese AM, Montazeri M, et al. Burden and Impact of Reactogenicity among Adults Receiving COVID-19 Vaccines in the United States and Canada: Results from a Prospective Observational Study. Vaccines (Basel). 2024;12(1).

31.	Walmart. Extra Strength Acetaminophen Tablets 500 mg, East Tabs, 24 tablets. [Available from: https://www.walmart.ca/en/ip/extra-strength-acetaminophen-tablets-500-mg/6000200614456?classType=REGULAR&athbdg=L1102&from=/search.

32.	Canadian Institute for Health Information. National Prescription Drug Utilization Information System — Plan Information Document, July 31, 2021. Ottawa, ON: CIHI;. 2021.

33.	Canadian Institute for Health Information (Ottawa O. Patient Cost Estimator Results. 2024.

34.	Sander B, Mishra S, Swayze S, Sahakyan Y, Duchen R, Quinn K, et al. Population-Based Matched Cohort Study of COVID-19 Healthcare Costs, Ontario, Canada. Emerging infectious diseases. 2025;31(4):710-9.

35.	Statistics Canada. Table 14-10-0327-01 Labour force characteristics by gender and detailed age group, annual DOI. [Available from: <https://doi.org/10.25318/1410032701-eng>.

36.	Statistics Canada. Table 14-10-0320-02  Average usual hours and wages by selected characteristics, monthly, unadjusted for seasonality (x 1,000). [Available from: <https://www150.statcan.gc.ca/t1/tbl1/en/tv.action?pid=1410032002>.
